# PREVALENCE AND ASSOCIATED FACTORS OF NOMOPHOBIA AMONG THE MEDICAL AND UNIVERSITY STUDENTS OF BANGLADESH

**DOI:** 10.64898/2026.01.10.26343847

**Authors:** Tonmoy Chowdhury, Saurya Dhungel, Mohammad Azmain Iktidar, Sreshtha Chowdhury, Purzia Tanaz Haque, Eshita Dey, Bikramaditya Chakma, Arefin Naher Oishee, Ishrat Jahan, Arpita Mazumder, Simanta Roy

## Abstract

**Background:** Nomophobia, characterised by the dread or anxiety of being without access to a mobile phone, has become an increasing behavioural issue among young adults, especially university students. Excessive reliance on smartphones has been linked to psychological suffering, impaired everyday functioning, and negative academic results. Nonetheless, information about the prevalence and determinants of nomophobia among university students in Bangladesh is scarce. This study aimed to evaluate the prevalence of nomophobia and investigate its associated sociodemographic traits and smartphone-related behaviors among medical and university students in Bangladesh.

**Materials and Methods:** A cross-sectional study was performed with 476 undergraduate medical and non-medical students from eight districts of Bangladesh from September 2023 to July 2025, using an online structured questionnaire. Nomophobia was evaluated via the validated Nomophobia Questionnaire (NMP-Q). The questionnaire was pilot-tested (n=60) and showed good reliability (Cronbach’s α=0.82). Information on socio-demographics, mobile phone usage trends, and application preferences was collected. Data was processed using STATA version 16. Descriptive statistics, t-tests, ANOVA, Pearson correlation, and multiple linear regression analyses were used to ascertain factors related to nomophobia.

**Results:** The average age of participants was 20.70 ± 1.52 years. 46.79% of students exhibited moderate nomophobia, whereas 25.69% had severe nomophobia. Elevated nomophobia scores were strongly correlated with female gender, moderate household income, usage of social media and communication applications, prolonged daily mobile phone usage (>7 hours), frequent phone checking, and instant phone checking upon awakening. Multiple linear regression indicated that extended phone usage, engagement with social communication applications, middle-income position, and early-morning phone checking are independent predictors of elevated nomophobia scores.

**Conclusion:** Nomophobia is significantly common among university students in Bangladesh. Behavioural patterns such as prolonged daily smartphone use, frequent phone checking, immediate phone use upon waking, and extensive engagement with social communication and media applications are associated with excessive smartphone usage and development of nomophobia. These findings underscore the necessity for awareness initiatives, early detection, and focused interventions to alleviate the adverse psychological and behavioural effects of nomophobia in students.

## Introduction

Nomophobia-short for ‘No Mobile Phone Phobia’ - refers to discomfort, apprehension, nervousness, or distress attributable to the inability to access a mobile phone[1,2]. In 2008, UK-based research institute introduced the term ‘Nomophobia’. They conducted a study which found that 53% of cell phone users exhibit anguish when they are faced with troubles regarding their phone [3]. Investigations reveal that Nomophobia is an emerging concern in the present times [4].

Mobile phones, an essential feature of our daily lives [5] have evolved into a multidimensional high-efficiency device that can be used as a media player, a tool for social networking, for playing video games and music, as a digital camera, for browsing [6] calls, texts, paying bills, and carrying out online transactions, among other functions [7]. This wide range of usage of phones makes it increasingly difficult to identify disorders like nomophobia [8]. Globally, the rate of subscription to cell phones is 103.5 per 100 population [9]. According to World Bank Trend statistics, in 2023, Bangladesh recorded over 114 mobile cellular subscriptions per 100 individuals, indicating an average of over one SIM card per person, attributable to multiple SIM ownership [10]. Published population health research indicates that subscription density rose from 0.22 per 100 individuals in 2000 to 107 by 2020 in Bangladesh [11]. Mobile phones create a perception of social inclusion among users [12]. Different studies done in India have found that several demographic variables, such as gender, age, frequency of use, and level of education, show significant relations to Nomophobia [8]. Specifically, women are more prone to Nomophobia than men, which is consistent with other studies that report a higher prevalence of Nomophobia among women [8,13–25]. Although no notable gender differences were found in smartphone use, nomophobia scores between men and women showed remarkable deviation [11,13,14,16–20,22,26–28]. These findings may be substantiated since women frequently fear losing social connections and want to evade loneliness in public settings [29]. The age group of 18 to 25 is reportedly susceptible to Nomophobia [8,20,30]. Still, the duration of using a mobile phone happens to be the strongest predictor of nomophobia, especially those who spend more than three hours on their device per day are susceptible to severe nomophobia [8,20,22,30]. Another study found a correlation between nomophobia score and anxiety and depression [1,21,30–35].

The findings of another study revealed the prevalence of nomophobia among medical college students and interns in Southern Haryana, India, was nearly 40% [36]. The study also reported significant association of nomophobia with sleep quality and academic activities. Other studies have shown that prevalence of moderate or severe nomophobia ranges from 50% to 85% [23–25,37–40]. Given the rising concern of nomophobia, this study aimed to evaluate the prevalence of nomophobia and investigate its associated sociodemographic traits and smartphone-related behaviours among medical and university students in Bangladesh.

## Methodology

### Study design and sampling technique

A cross-sectional study was carried out among undergraduate students from eight districts in Bangladesh -Dhaka, Chattogram, Rajshahi, Khulna, Sylhet, Barisal, Mymensingh, and Rangpur. Participants were recruited through snowball sampling.

### Study Instrument and participants

The data was collected from the study participants using a structured online questionnaire between September 2023 to July 2025. Since the study’s major goal was to examine the determinants and prevalence of nomophobia, it included only adult (over 18 years of age) students in its eligibility evaluation. Students who were non-citizen of Bangladesh or did not own a mobile device were excluded from the study. Prior to the main study, the questionnaire underwent a pilot test among 60 participants to ensure its reliability using Cronbach’s alpha (α=0.82).

### Independent variables

Information about the respondent’s socioeconomic status (age, sex, occupation, and place of employment), type of digital device used (Android, iPhone, Desktop, laptop, tablet, etc.), type of application or social media used (Facebook, What’s App, or any streaming media), and self-reported average duration of device use (2 hours, 2-4 hours, 5-7 hours, or >7 hours) before and after going to sleep were assessed. All participants who provided their consent to be included before their data were collected.

### Dependent variables

For assessing nomophobia, we used the NMP-Q questionnaire [41]. It is a 20-item scale that is scored based on a seven-point Likert scale, where 1 indicates ‘does not agree at all’ and 7 indicates ‘strongly agree’. Higher scores indicate higher levels of nomophobia. A study conducted in adolescents and young university adults also used the Nomophobia Questionnaire (NMP-Q) instrument and the standard cut-off points [13], where a score of 20 indicates absence; scores between 21 and 59 indicates mild level; scores between 60 and 99 indicate moderate level; and scores of 100 or above indicate severe level [42] Furthermore, 1-60, 61-100, 101-140 indicated mild, moderate, and severe nomophobia, respectively [23] [42].

### Statistical analysis

STATA, version 16, was used to analyze the data. The quantitative data was summarized using the mean and the standard deviation. The frequency of occurrence of a categorical variable was stated as a percentage. We used the t-test and Anova to look at the association between categorical variables with mean NMP-Q scores, and multiple linear regression to assess the predictors of nomophobia.

### Ethical consideration

The Institutional Review Board of Public Health Foundation Bangladesh approved the research (Approval no: PHFBD-ERC-IP09/2023), and all participants provided informed written consent. Wherever feasible, the 1964 Declaration of Helsinki and later modifications, latest on October 2024 and comparable ethical standards were followed. Data collection was voluntary, and no incentives were offered to participants. Data was only accessible to the research team and were not disclosed anywhere.

## Results

The study involved 476 participants, with a mean age of (20.70±1.52) years and a mean BMI of (22±3.88) kg/m^2^(Table 1). The majority of the participants were enrolled in medical school (53.4%), female (53%), and from Chattogram (46.3%). Additionally, a significant portion of the participants (54.2%) lived away from their families. Most participants (57.3%) came from well-off families whose monthly income was 30000-60000 Bangladeshi Taka (BDT) [43–45]. Among the participants, android smartphones were the most commonly used device (92.7%), while a smaller proportion used laptops (30.7%). Other devices used included button phones (6.2%), iPhones (9.9%), desktops (6.4%), and tablets (4.8%). The majority of participants accessed the internet via Wireless Fidelity (Wi-Fi) (75.5%), while a smaller portion used mobile/cellular data (22.7%). Social media applications (e.g., Facebook, Instagram) were predominantly used by participants (73.6%), followed by social communication applications (e.g., WhatsApp, Messenger; 67.9%). A notable proportion (49.3%) indicated an inclination towards using music and video sharing applications (e.g., YouTube, Netflix, Spotify, TikTok). A similar picture was portrayed when the purpose of their mobile phone use was asked. The majority of the participants (82.6%) used it to communicate with family and friends. Almost equal numbers of participants used their mobile phones for browsing and sharing on social media, and to follow the news and current events. The majority (41.5%) of all participants used their phones daily for 2-4 hours. A large majority (94.5%) used their phones before going to bed, and upon waking up, most of them (78.4%) immediately checked their devices (Table 1).

**Table 1:**
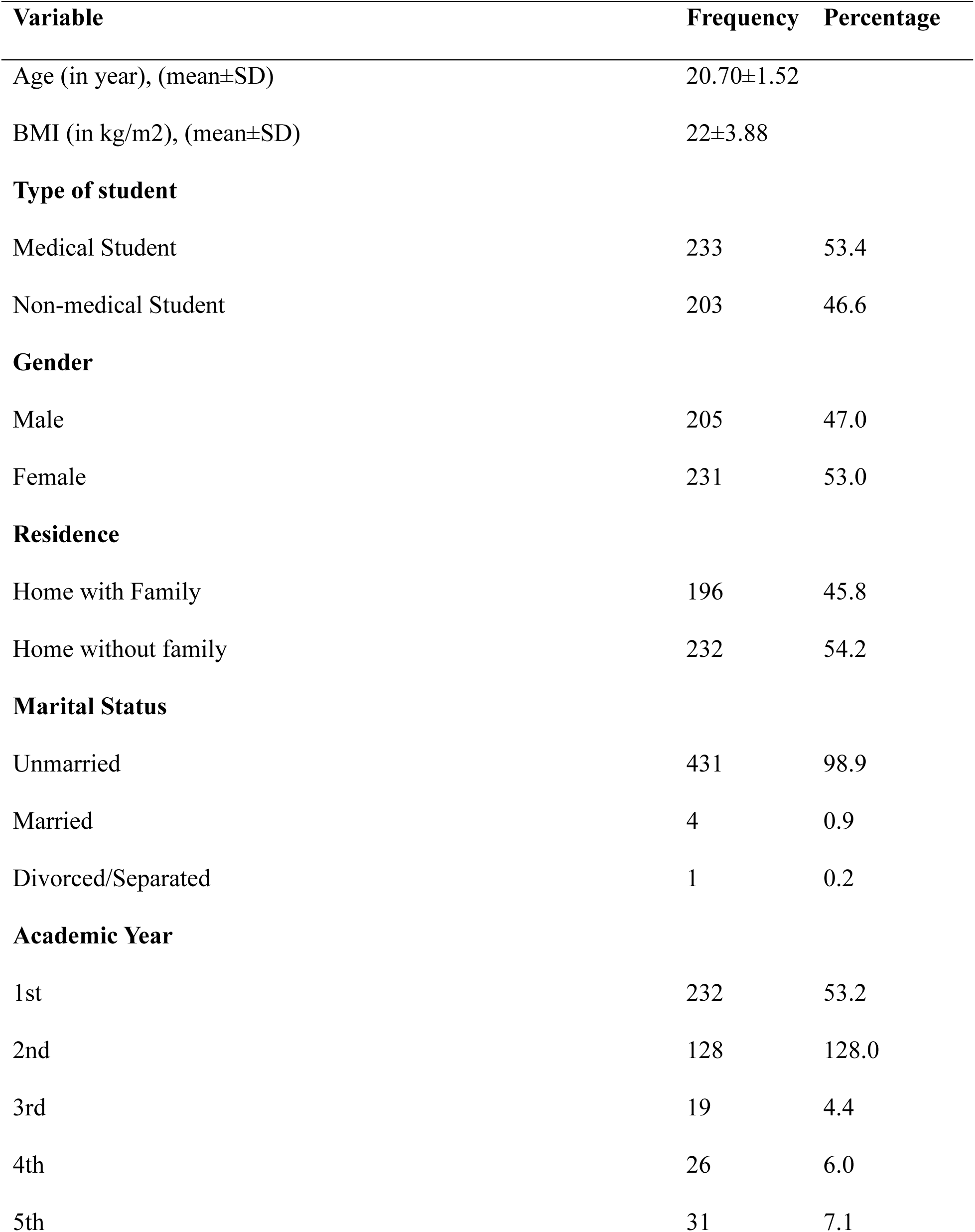
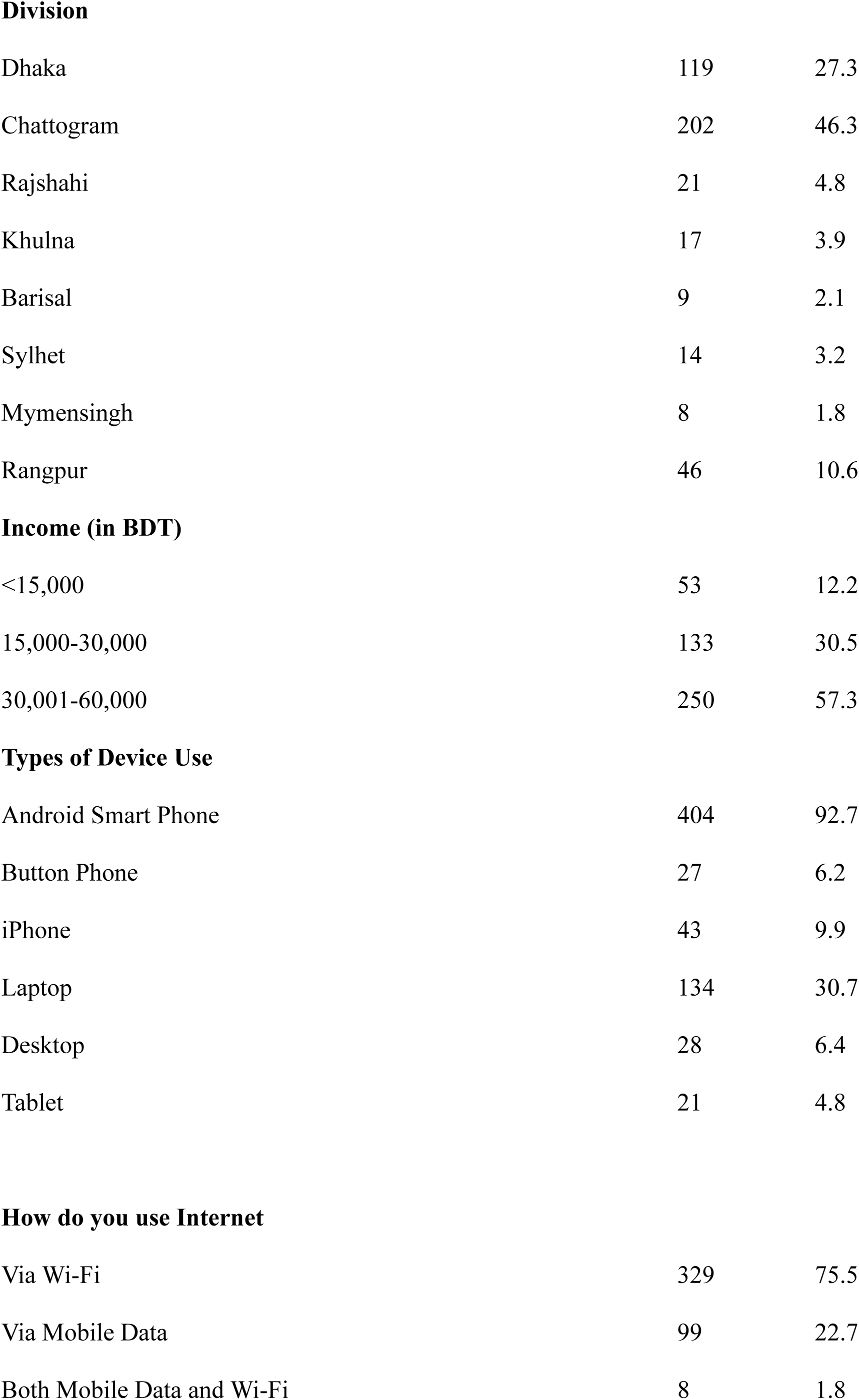
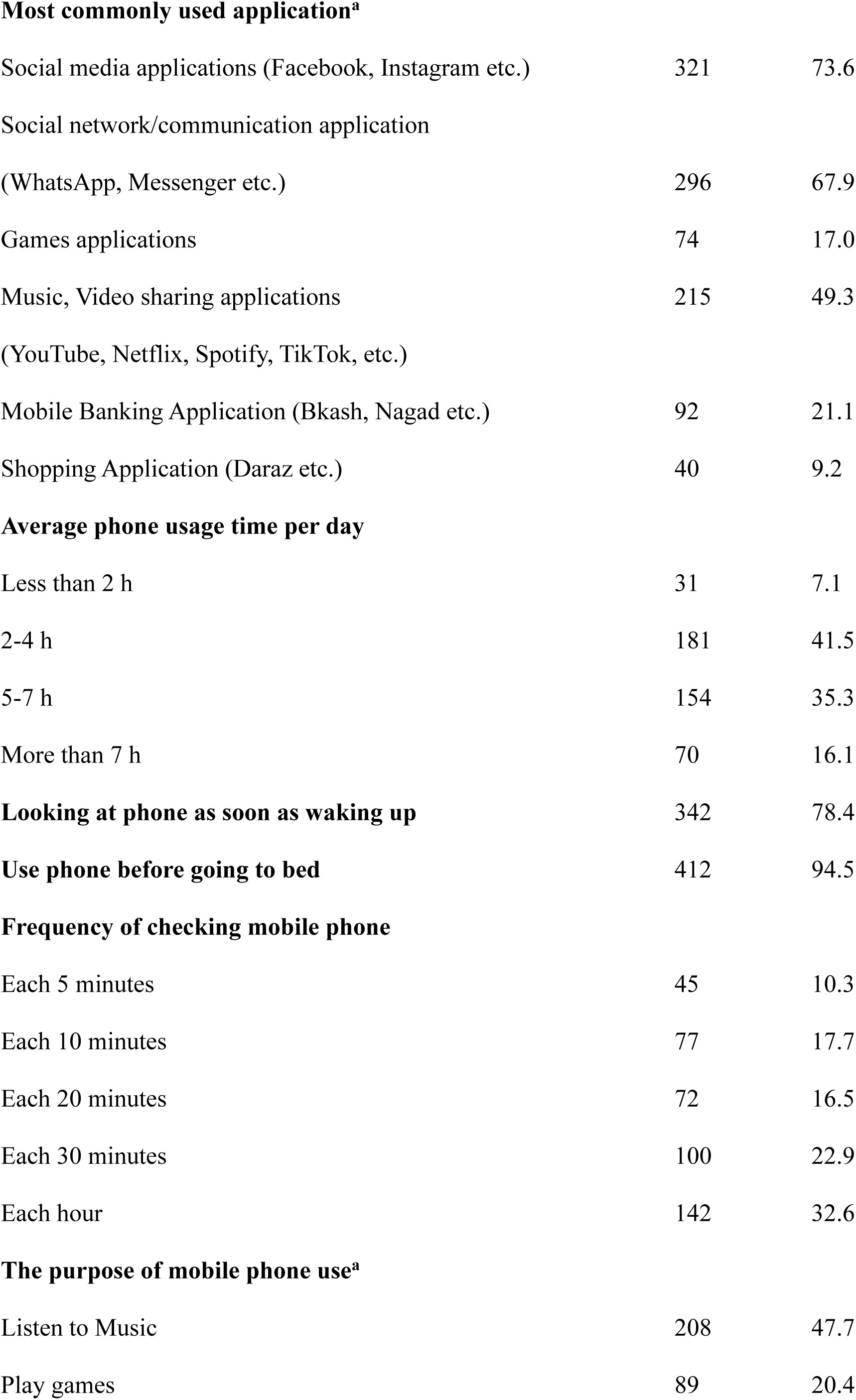
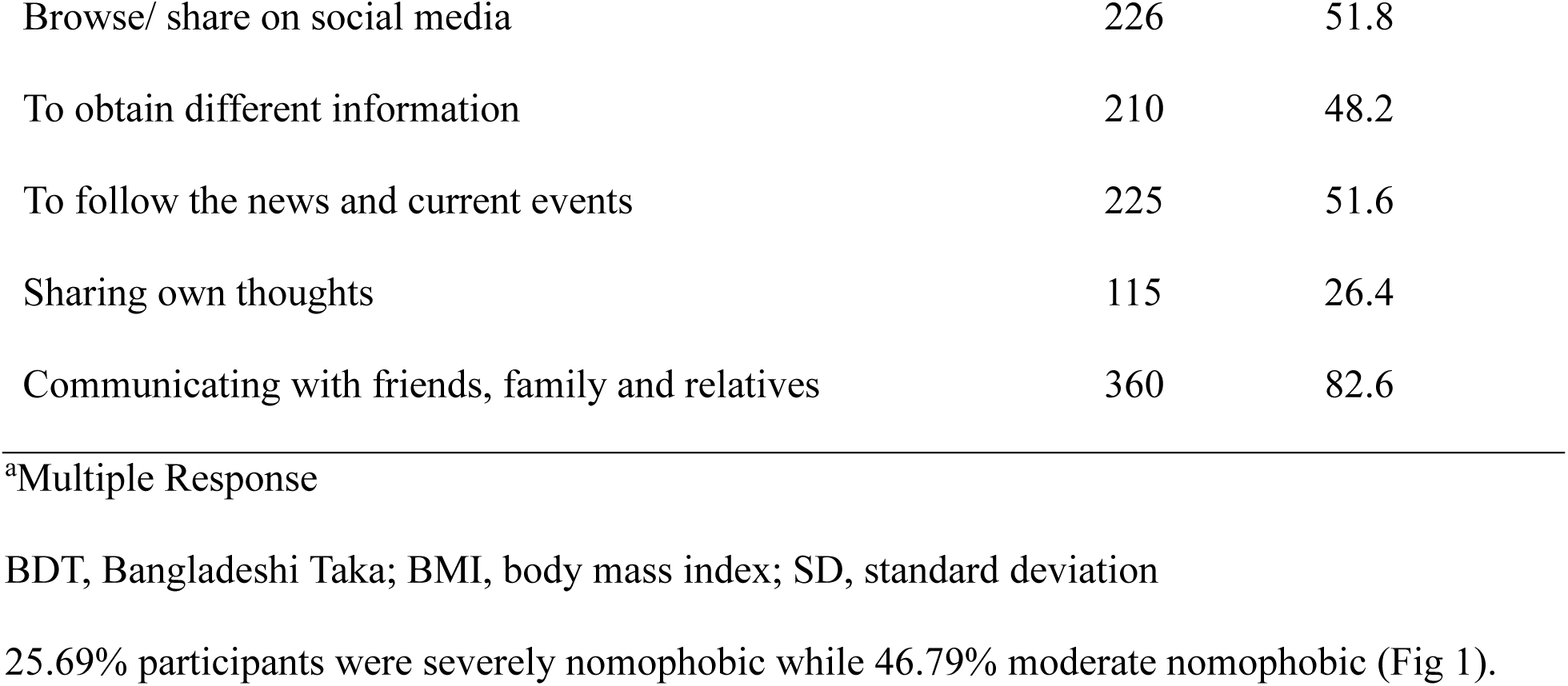
Background Characteristics of Study Participants (n=436)

| Variable | Frequency | Percentage |
| --- | --- | --- |
| Age (in year), (mean±SD) | 20.70±1.52 |  |
| BMI (in kg/m2), (mean±SD) | 22±3.88 |  |
| **Type of student** |  |  |
| Medical Student | 233 | 53.4 |
| Non-medical Student | 203 | 46.6 |
| **Gender** |  |  |
| Male | 205 | 47.0 |
| Female | 231 | 53.0 |
| **Residence** |  |  |
| Home with Family | 196 | 45.8 |
| Home without family | 232 | 54.2 |
| **Marital Status** |  |  |
| Unmarried | 431 | 98.9 |
| Married | 4 | 0.9 |
| Divorced/Separated | 1 | 0.2 |
| **Academic Year** |  |  |
| 1st | 232 | 53.2 |
| 2nd | 128 | 128.0 |
| 3rd | 19 | 4.4 |
| 4th | 26 | 6.0 |
| 5th | 31 | 7.1 |
| **Division** |  |  |
| Dhaka | 119 | 27.3 |
| Chattogram | 202 | 46.3 |
| Rajshahi | 21 | 4.8 |
| Khulna | 17 | 3.9 |
| Barisal | 9 | 2.1 |
| Sylhet | 14 | 3.2 |
| Mymensingh | 8 | 1.8 |
| Rangpur | 46 | 10.6 |
| **Income (in BDT)** |  |  |
| <15,000 | 53 | 12.2 |
| 15,000-30,000 | 133 | 30.5 |
| 30,001-60,000 | 250 | 57.3 |
| **Types of Device Use** |  |  |
| Android Smart Phone | 404 | 92.7 |
| Button Phone | 27 | 6.2 |
| iPhone | 43 | 9.9 |
| Laptop | 134 | 30.7 |
| Desktop | 28 | 6.4 |
| Tablet | 21 | 4.8 |
| **How do you use Internet** |  |  |
| Via Wi-Fi | 329 | 75.5 |
| Via Mobile Data | 99 | 22.7 |
| Both Mobile Data and Wi-Fi | 8 | 1.8 |
| **Most commonly used application<sup>a</sup>** |  |  |
| Social media applications (Facebook, Instagram etc.) | 321 | 73.6 |
| Social network/communication application<br>(WhatsApp, Messenger etc.) | 296 | 67.9 |
| Games applications | 74 | 17.0 |
| Music, Video sharing applications<br>(YouTube, Netflix, Spotify, TikTok, etc.) | 215 | 49.3 |
| Mobile Banking Application (Bkash, Nagad etc.) | 92 | 21.1 |
| Shopping Application (Daraz etc.) | 40 | 9.2 |
| **Average phone usage time per day** |  |  |
| Less than 2 h | 31 | 7.1 |
| 2-4 h | 181 | 41.5 |
| 5-7 h | 154 | 35.3 |
| More than 7 h | 70 | 16.1 |
| **Looking at phone as soon as waking up** | 342 | 78.4 |
| **Use phone before going to bed** | 412 | 94.5 |
| **Frequency of checking mobile phone** |  |  |
| Each 5 minutes | 45 | 10.3 |
| Each 10 minutes | 77 | 17.7 |
| Each 20 minutes | 72 | 16.5 |
| Each 30 minutes | 100 | 22.9 |
| Each hour | 142 | 32.6 |
| **The purpose of mobile phone use<sup>a</sup>** |  |  |
| Listen to Music | 208 | 47.7 |
| Play games | 89 | 20.4 |
| Browse/ share on social media | 226 | 51.8 |
| To obtain different information | 210 | 48.2 |
| To follow the news and current events | 225 | 51.6 |
| Sharing own thoughts | 115 | 26.4 |
| Communicating with friends, family and relatives | 360 | 82.6 |

<sup>a</sup>Multiple Response
BDT, Bangladeshi Taka; BMI, body mass index; SD, standard deviation

Among the students, factors such as gender (p-value = 0.018), income in BDT (p-value = 0.044), social media applications use (p-value = 0.014), social communication application use (p-value = 0.005), music, video sharing application use (p-value = 0.002), mobile banking application use (p-value = 0.036) and shopping application use (p-value = 0.040) had significant relation with having higher nomophobia (Table 2). Average phone usage per day (p-value = 0.022), looking at phone as soon as waking up (p-value = 0.010) and frequency of checking mobile phone (p-value = 0.006) had a significant association with developing nomophobia (Table 2).

**Table 2:**
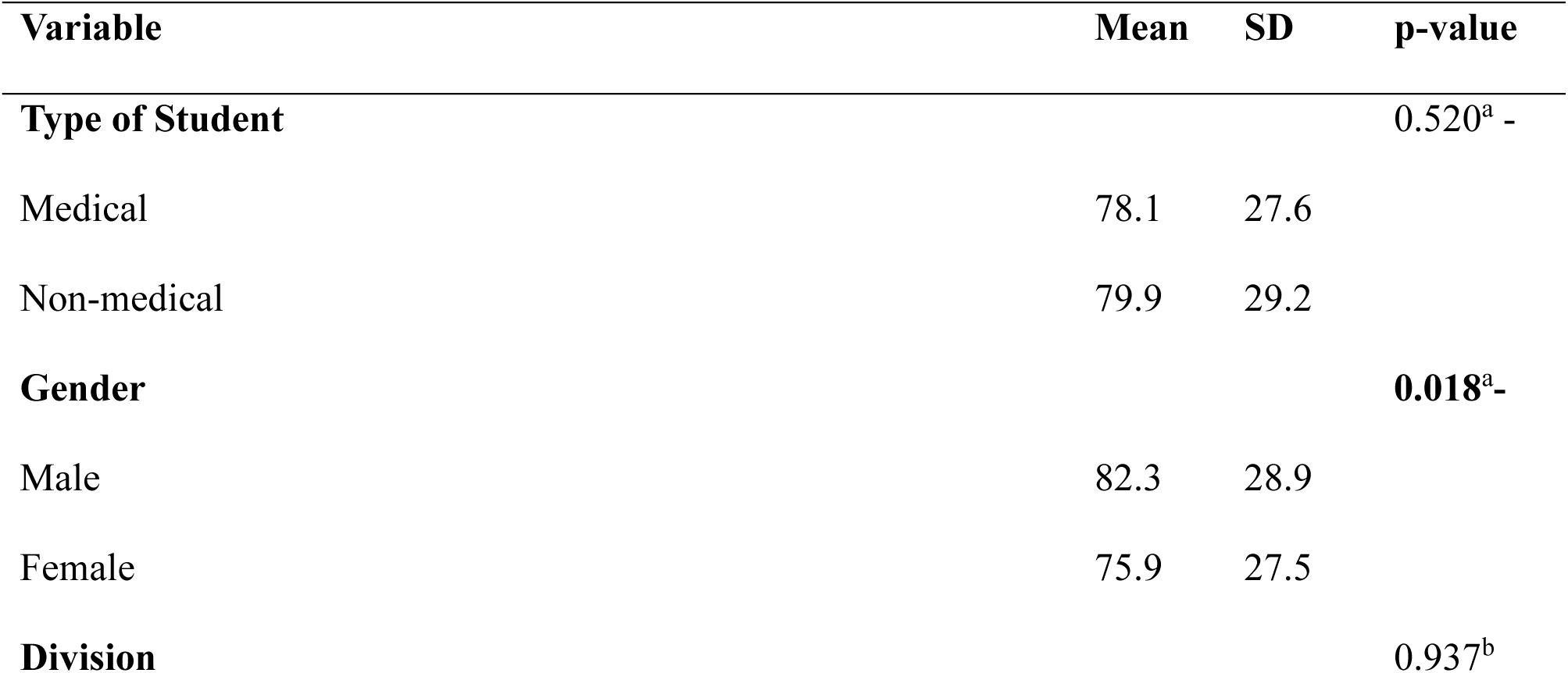
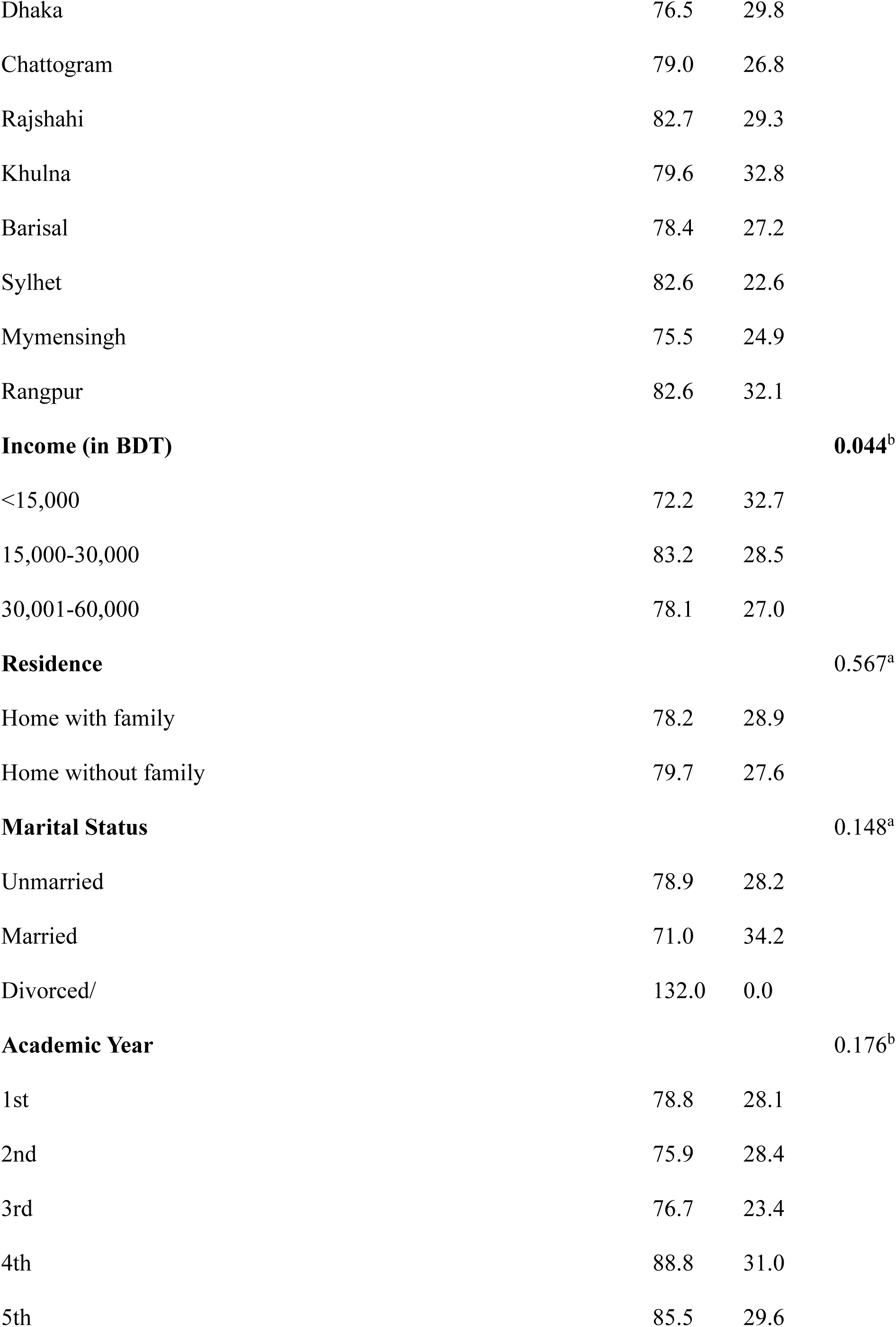
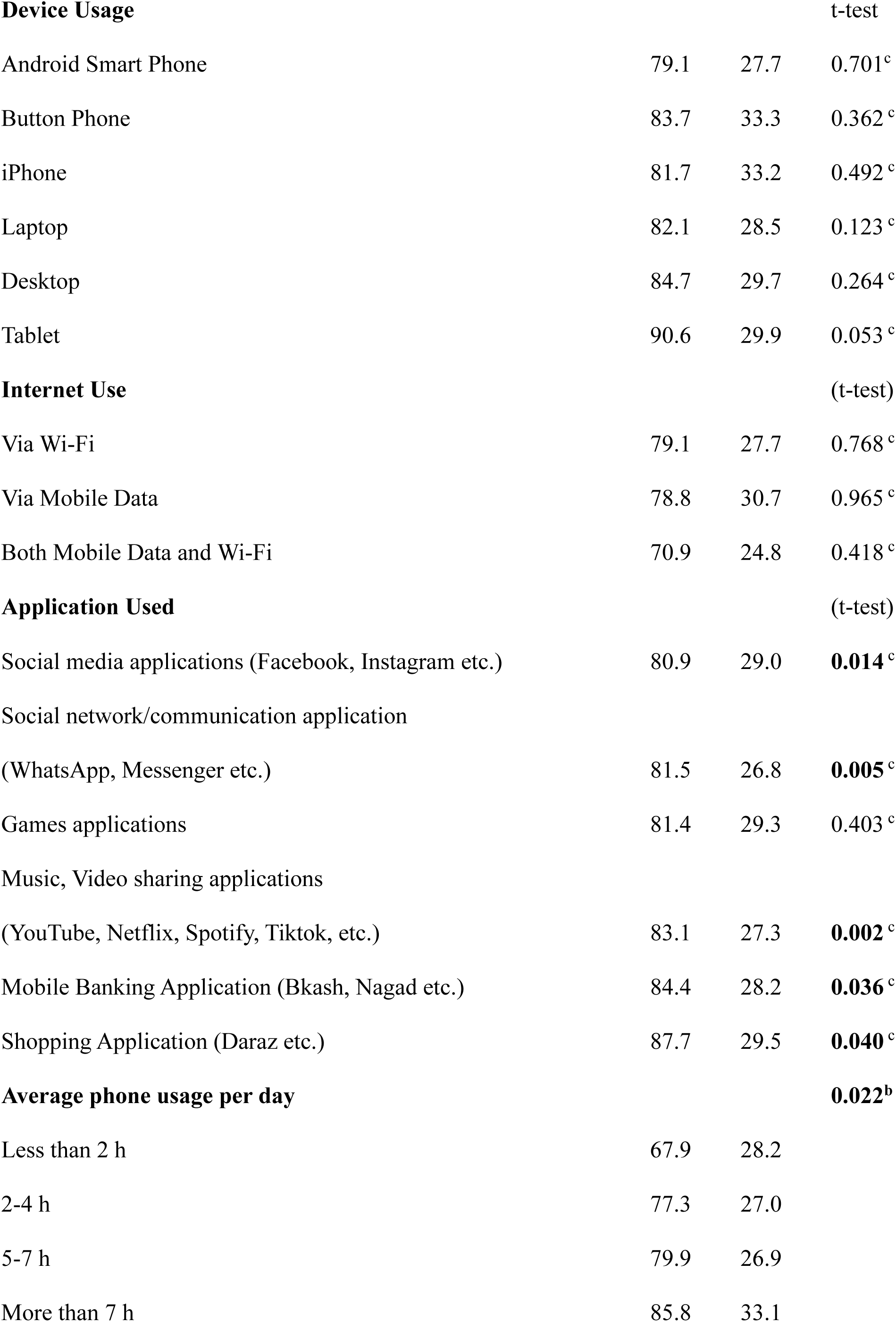
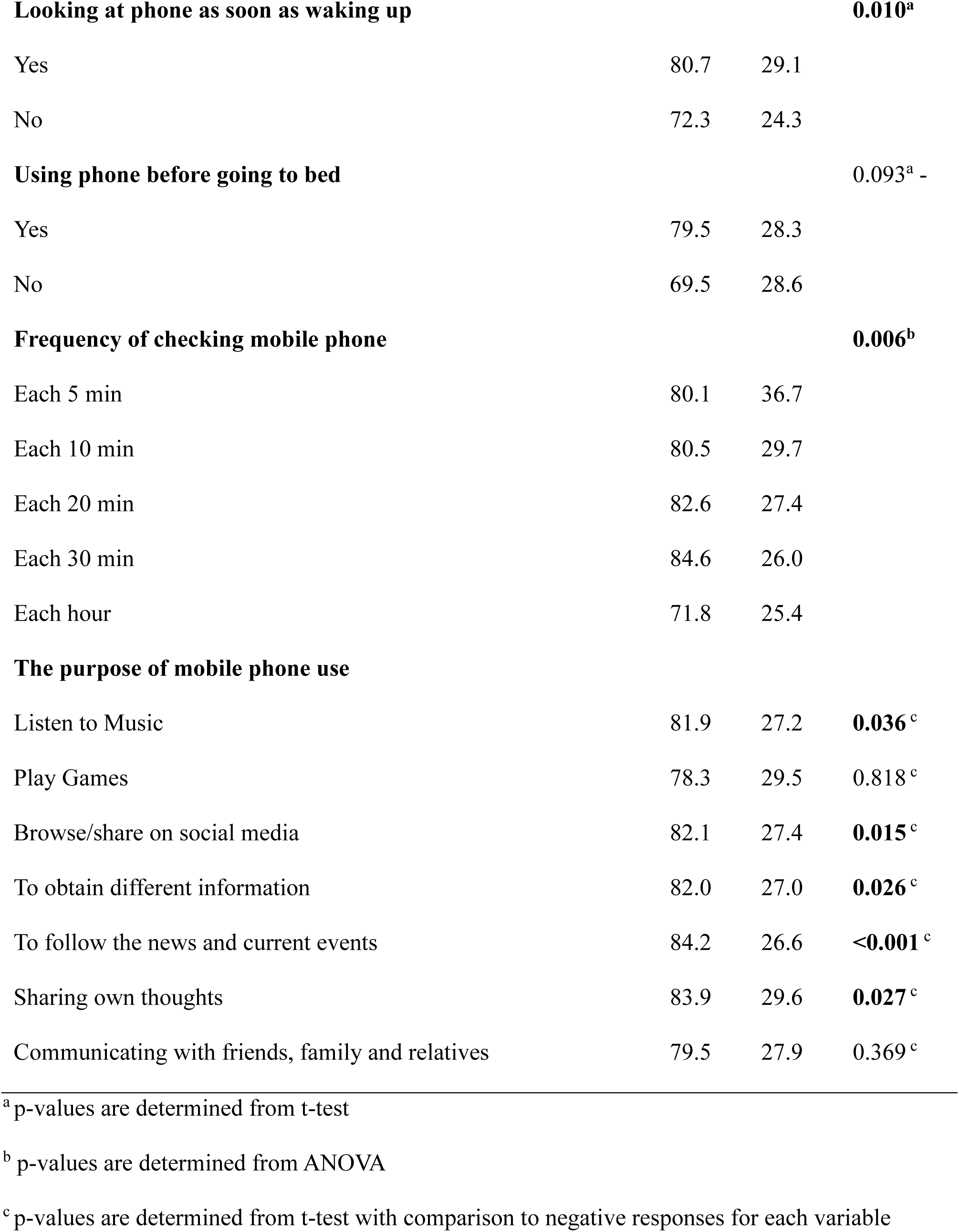
Bivariate relationship between the participants’ characteristics and NMP-Q total score (n=436)

**Table 3:** Multiple linear regression for the predictors of NMP-Q total score.

| Variables | $\beta$ | 95% CI | p-value |
| --- | --- | --- | --- |
| **Gender** |  |  |  |
| Male | Ref. |  |  |
| Female | -5.24 | (-10.57, 0.10) | 0.054 |
| **Income (in BDT)** |  |  |  |
| <15,000 | Ref. |  |  |
| 15,000-30,000 | 11.48 | (2.57, 20.39) | **0.012** |
| 30,001-60,000 | 6.74 | (-1.58, 15.07) | 0.112 |
| **Social network/communication application (i.e., WhatsApp, Messenger etc.) use** |  |  |  |
| No | Ref. |  |  |
| Yes | 8.25 | (2.63, 13.86) | **0.004** |
| **Average phone usage per day** |  |  |  |
| Less than 2 h | Ref. |  |  |
| 2-4 h | 4.71 | (-6.01, 15.42) | 0.388 |
| 5-7 h | 5.66 | (-5.37, 16.69) | 0.314 |
| More than 7 h | 13.20 | (1.31, 25.10) | **0.030** |
| **Looking at phone as soon as waking up** |  |  |  |
| No | Ref. |  |  |
| Yes | 7.09 | (0.67, 13.52) | **0.031** |

### Significant (<0.05) p-values are in bold

We also calculated the Pearson correlation coefficient to assess the relation between nomophobia scores with age and body mass index (BMI) [data not provided in table]. Age had a statistically significant, although a weak positive relation with nomophobia scores (r = 0.12; p-value = 0.015). Moreover, BMI did not have any significant correlation with nomophobia scores (r = 0.04; p-value = 0.44)

Participants who had an income between 15,000 to 30,000 BDT had on average, 11.48 units higher nomophobia scores (β=11.48; 95% CI: 2.57, 20.39; p=0.012) than those who earned less than 15,000 BDT, after adjusting for gender, use of communication application, average phone usage per day, and looking at the phone as soon as waking up (Table43). Furthermore, people using social media had on average 8.25 units higher nomophobia scores (β=8.25; 95% CI: 2.63, 13.86; p=0.004) than those who were not using any social media platforms, after adjusting for gender, income, average phone usage per day, looking at the phone as soon as waking up. Participants using mobile phones for more than 7 hours had on average, 13.20 units higher nomophobia scores (β=13.20; 95% CI: 1.31, 25.10; p=0.030) than those who used mobile phone for less than 2 hours, after adjusting for gender, income, use of communication applications, and looking at the phone as soon as waking up. Additionally, participants who looked at their mobile phones as soon as they woke up had on average, 7.09 units higher nomophobia scores (β=7.09; 95% CI: 0.67, 13.52; p=0.031) than those who did not, after adjusting for gender, income, use of communication applications, and average phone usage per day.

## Discussion

The study revealed a significant prevalence of nomophobia among university students in Bangladesh, with approximately 73% exhibiting moderate to severe symptoms, highlighting a considerable behavioural issue. Female students and those from middle-income households exhibited markedly elevated nomophobia ratings, underscoring the impact of demographic and socioeconomic variables. Extended daily mobile phone usage (>7 hours), and instant checking of phone upon awakening were significantly correlated with elevated nomophobia levels. Usage of social media and communication platforms was substantially associated with elevated nomophobia scores.

In our study 53.4% of the total participants were medical students and most of the participants were more addicted to mobile phones. A similar cross-sectional study conducted in India concluded that all medical students had various degrees of nomophobia which is really concerning [46]. A cross-sectional study was carried out in March, 2021 in Pakistan which led to the conclusion that nomophobia, in mild to severe forms, affects the majority of undergraduate students in Pakistan [47]. Our study also depicted that among the participants, 26.15% were mildly nomophobic, 46.79% were moderately nomophobic and 25.69% were severely nomophobic. Furthermore, a study done in November 2019 in India shows that a large fraction of the students displayed severe nomophobia symptoms, distinct usage habits, and false beliefs about their health and usage pattern [8]. In a recent study done on Bangladeshi young adults, 61.4% had higher Problematic Use of Mobile Phone (PUMP) scores [48–50]. The Problematic Use of Mobile Phone (PUMP) scale is a validated self-reported tool designed to evaluate maladaptive, excessive, and dependency-like behaviours associated with mobile phone usage and their resultant effects [51]. Other studies in different countries conducted with NMP-Q showed results ranging from 42.6% to 67.2% with moderate or severe nomophobia [21,42,52,53].

In our study, we found a positive correlation between different ages and nomophobia. Similar findings were present in a study conducted in North India [21]. A study done in August 2020 found that nomophobia is a recent mental health issue, particularly in male teens [54]. On the contrary, our study showed that females were more prone to be nomophobic. Two separate studies conducted in Saudi Arabia and Turkey observed no correlation between age and nomophobia [42,48]. Being female had significant association with nomophobia in our study and the study conducted in Turkey[55]. Our study depicted that females (53%) were prone to be more nomophobic which was similar to the study published in Turkey [55]. Our study reveals the multi-purpose usage of mobile phones by students. Social networking, mobile banking, shopping and video sharing are the significant purposes of using phones that have association with nomophobia, similar to other studies[15,56]. In our study, there is a statistically significant relationship between the number of hours per day spent using a mobile phone and the severity of nomophobia. Those who use more than 7 hours of mobile phone had higher nomophobia scores. A similar association between prolonged daily phone use and nomophobia was exhibited in other studies [21,48]. Another study observed 62.1% of students spent time on smartphone for more than 3 hours [21]. In our study, 78.4% of students practice the habit of checking their phones as soon as they wake up, which has a strong correlation with the development of nomophobia. A similar association between instant checking of phone after waking up was observed by Hoşgör [57]. A similar association between nomophobia and frequency of checking mobile phones was observed by Khilnani [58]. According to a study conducted in Peru, nomophobia is a common and rising issue among college students, which manifests earlier in life and is linked to symptoms of anxiety or depression [59]. The mental health of the students might benefit from the use of evaluation and early intervention measures [32].

This study possesses certain limitations that must be acknowledged when evaluating the results. The cross-sectional design limits the capacity to determine causal correlations between nomophobia and its related factors. The detected linkages indicate correlations at a specific moment and cannot ascertain directionality or temporal succession. The study utilised self-reported data obtained via an online questionnaire, which may be influenced by recall bias and social desirability bias. Participants may have inaccurately reported their mobile phone usage patterns and nomophobia-related behaviours. Third, the implementation of snowball sampling and online data collecting may have resulted in selection bias, as students with consistent internet access and greater engagement with mobile devices were more inclined to participate. Thus, the sample may not comprehensively reflect all university students in Bangladesh, especially those from rural regions or lower socioeconomic strata. Fourth, significant psychological and behavioural confounders, including anxiety, depression, stress levels, sleep quality, and academic pressure, were not directly evaluated, despite existing evidence indicating correlations between them and nomophobia. The lack of information in these variables may have resulted in residual confounding in the regression analysis. Ultimately, although the NMP-Q is a validated and extensively utilised tool, it assesses perceived nomophobia rather than clinically recognised behavioural addiction. Consequently, the results should be understood as indicative of nomophobia symptomatology rather than clinical illness. Despite the limitations, this study is important because understanding the prevalence and associated factors of nomophobia among mobile phone users can help in developing strategies to prevent its negative effects, such as discomfort, anger, anxiety, and feelings of insecurity [40].

### Conclusion

In a nutshell, nomophobia is a pressing issue in today’s world. In this modern era, avoiding smartphones and devices is nearly impossible as we are hugely dependent on these devices in our day-to-day activities. Therefore, it is crucial to raise awareness of the negative impacts of nomophobia and conduct extensive research on it.

## Data Availability

All relevant data are within the manuscript and its Supporting Information files.

## Acknowledgement

The authors express their profound gratitude to all study participants for their voluntary involvement. The authors acknowledge the significant assistance and support rendered by Dipta Das Tirtha, Abu Talib Shakib, and Antara Das Gupta during different phases of the study process. The authors express gratitude to all people who helped, directly or indirectly, to the successful completion of this study.

## Supporting Information

**S1 File. Dataset**

**S2 File. Fig 1: Prevalence of nomophobia among the study participants**

